# Structural network topology correlates with cognitive impairment in Parkinson’s disease

**DOI:** 10.1101/2023.09.18.23295719

**Authors:** Zhichun Chen, Guanglu Li, Liche Zhou, Lina Zhang, Li Yang, Jun Liu

## Abstract

In patients with Parkinson’s disease (PD), cognitive impairment is common and substantially exacerbates disease severity. The objective of this study is to investigate whether cognitive decline affects disease severity and whether structural network disruption contributes to cognitive impairment in PD patients. PD patients (n = 146) receiving structural and functional magnetic resonance imaging from Parkinson’s Progression Markers Initiative (PPMI) database were included. Based on the scores of Symbol Digit Modalities Test (SDMT), patients were classified into lower quartile group (SDMT score rank: 0%∼25%), interquartile group (SDMT score rank: 26%∼75%), and upper quartile group (SDMT score rank: 76%∼100%) according to their SDMT score quartiles to assess how cognitive function shapes disease manifestations and brain network topology. Patients in the lower quartile group exhibited more severe motor symptoms, non-motor symptoms, and striatal dopamine depletion compared to interquartile group and upper quartile group. In addition, they also showed impaired structural network topology both at global and local levels compared to interquartile group and upper quartile group. Furthermore, different nodal network metrics in structural network contributed to cognitive and motor impairment of PD patients. Taken together, PD patients with worse cognitive function exhibited more severe motor and non-motor symptoms and aberrant structural network metrics were associated with both cognitive and motor decline of PD patients.

## 1. Introduction

In addition to its typical motor symptoms, Parkinson’s disease (PD) is associated with a variety of non-motor manifestations that prominently increase the overall disease burden. Cognitive decline is almost 6 times more frequent in PD patients than in the healthy people and is one of the key non-motor features [1]. It can occur at any disease stage, even at the prodromal phase and early stage of the disease [2, 3]. It is associated with faster disease progression and worse prognosis [1, 4], which draws plenty of attention to promote the early diagnosis and treatment of cognitive impairment during PD management. According to previous literature, PD patients can display the impairments of multiple cognitive domains, spanning episodic memory [5], verbal memory [6], executive function [7], visuospatial ability [8], and processing speed [9]. Cognitive impairment is associated with multiple risk factors, including age, education, hypertension, motor severity, depression, anxiety, and excessive daytime sleepiness (EDS) [10–13]. Although tremendous studies have focused on cognitive impairment in PD in past decades, the neural mechanisms underlying cognitive decline in PD patients remain largely elusive.

Recently, with multi-modal network analysis in functional magnetic resonance imaging (fMRI), researchers have identified widespread changes of structural and functional networks in PD patients with cognitive impairment [14–16]. For structural network, PD patients display extensive disruptions of white matter integrity [17], which is associated with worse disease severity and cognitive decline [18, 19]. Particularly, the burden of white matter hyperintensity has been demonstrated to be an imaging biomarker of cognitive impairment in PD patients [20]. Therefore, white matter disruptions have been considered as an essential culprit for cognitive decline in PD patients. Consistently, we recently revealed that PD patients exhibited age-dependent destructions in multiple white matter tracts, which were significantly associated with cognitive impairment in PD patients [13]. The impairments of white matter tracts induced dramatical changes of structural network topology in PD patients, which was also associated with cognitive decline in PD patients [6, 13]. Recently, we revealed that small-world properties in structural network were significantly associated with verbal memory decline in PD patients [6]. Additionally, we also demonstrated that structural topological properties, such as global efficiency and local efficiency partially mediated the age-dependent cognitive decline in PD patients [13]. Taken together, structural network aberrations may play an essential role in the development of cognitive decline in PD patients.

Although we have observed significant correlations between cognitive impairment and global network topology [6, 13], whether local network topology also contributes to cognitive impairment in PD remains largely unknown. To better investigate the associations between structural network topology and cognitive function in PD patients, we divide the PD patients into 3 cognition quartile groups according to their cognitive function measured by Symbol Digit Modalities Test (SDMT) [21]: lower quartile group (Q1, SDMT score rank: 0%∼25%), interquartile group (Q2–3, SDMT score rank: 26%∼75%), and upper quartile group (Q4, SDMT score rank: 76%∼100%). In consequence, the objective of this study is to examine whether structural topological metrics are significantly different among different cognition quartile groups and explore the associations between structural topological metrics and cognitive function in PD patients. The similar approaches for subgroup divisions based on quartiles of continuous variables have been described previously [13, 22, 23]. Specifically, our research goals include: (i) to compare clinical assessments among different cognition quartile groups; (ii) to compare structural network metrics among different cognition quartile groups; (iii) to explore the associations between network metrics and clinical assessments, especially cognitive function; (iv) to examine whether differential network properties contribute to cognitive impairment using mediation analysis.

## 2. Materials and Methods

### 2.1. Participants

Data analyzed in this study were downloaded from Parkinson’s Progression Markers Initiative (PPMI) database, which is an international multicenter cohort study [24]. For currently updated information on this study, please visit www.ppmi-info.org. This dataset from PPMI included over 400 patients who were diagnosed with early PD, as well as 200 healthy participants who served as control. They went through standardized assessments, which included imaging, biochemical tests, clinical examinations, and behavioral evaluations. The purpose of these assessments was to promote the investigations of predictive biomarkers for PD progression. The procedures of PPMI study have been approved by the Institutional Review boards of each participating center, and all patients signed informed consents before participating the study. The inclusion criteria for PD patients consisted of: (i) being above the age of 30; (ii) being diagnosed with PD according to the MDS Clinical Diagnostic Criteria for PD; and (iii) undergoing 3D T1-weighted MPRAGE imaging and diffusion tensor imaging (DTI) within the same timeframe. Those participants who showed clear abnormalities in T1-weighted or T2-weighted MRI scans, or had genetic mutations linked to familial PD, or belonged to the genetic PPMI cohort and prodromal cohort were excluded from this study. Based on the criteria mentioned above, a total of 146 PD patients were included in the analysis. They underwent motor evaluations which consisted of Hoehn and Yahr stages, Tremor scores, Total Rigidity scores, and Movement Disorder Society Unified Parkinson’s Disease Rating Scale part III (MDS-UPDRS-III). The non-motor evaluations included Epworth Sleepiness Scale (ESS), REM Sleep Behavior Disorder Screening Questionnaire (RBDSQ), 15-item Geriatric Depression Scale (GDS), Scale for Outcomes in Parkinson’s Disease-Autonomic (SCOPA-AUT), State-Trait Anxiety Inventory (STAI), Semantic Fluency Test (SFT), Benton Judgment of Line Orientation test (BJLOT), Letter Number Sequencing test (LNS), Montreal Cognitive Assessment (MoCA), Hopkins Verbal Learning Test-Revised (HVLT-R) and SDMT. The patients also received ^123^I-ioflupane single-photon emission computed tomography (SPECT) scanning, which was performed in accordance with the technical instructions of the PPMI study (http://ppmi-info.org/). The striatal binding ratios (SBRs) for striatum and its subregions were computed from SPECT scans. Specifically, the SBRs were calculated based on the formula: (target region/reference region)-1, in which the reference region was occipital lobe. In order to evaluate the associations between cognitive function and clinical features or brain networks, PD participants (n = 146) were classified into lower quartile group (Q1, SDMT score rank: 0%∼25%, SDMT score range: 7∼34), interquartile group (Q2-3, SDMT score rank: 26%∼75%, SDMT score range: 34∼48), and upper quartile group (Q4, SDMT score rank: 76%∼100%, SDMT score range: 48∼76) based on their SDMT score quartiles. The clinical characteristics of PD patients in 3 quartile groups were shown in Table S1, Figure 1, and Figure S1. The clinical characteristics and selection criteria of 189 age– and sex-matched control participants have been reported in our recent study [13]. Because most of control participants had no MRI data, they were not included in the imaging analysis.

**Fig. 1.**
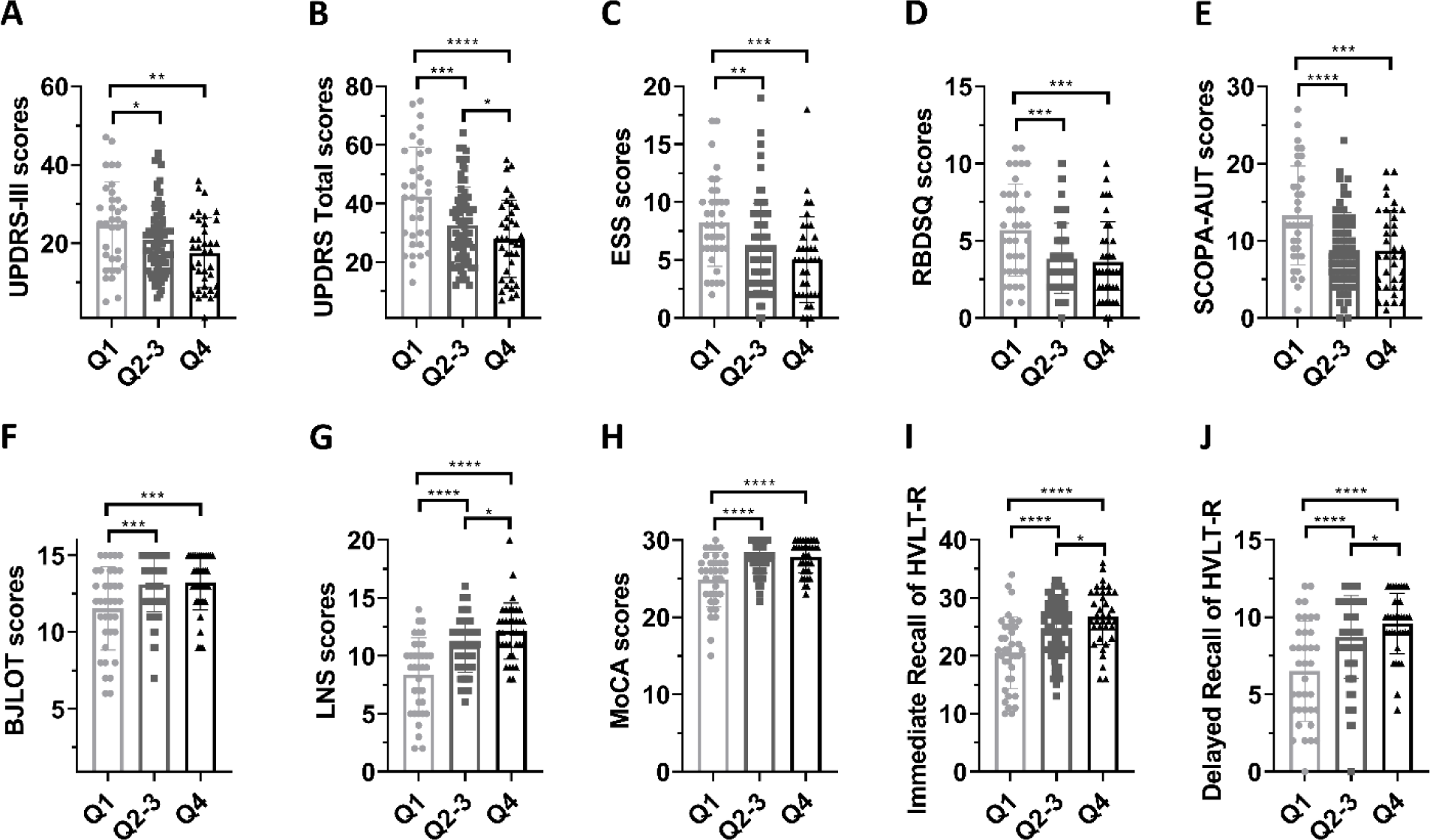
Group differences in clinical characteristics. (A-J) Group differences in scores of UPDRS-III (A), total UPDRS (B), ESS (C), RBDSQ (D), SCOPA-AUT (E), BJLOT (F), LNS (G), MoCA (H), Immediate Recall (I) and Delayed Recall (J) of HVLT-R among 3 quartile groups. One-way ANOVA followed by Tukey’s post hoc test (Q1 group *vs* Q2-3 group *vs* Q4 group) were conducted to compare clinical variables. *p* < 0.05 was considered statistically significant. \**p* < 0.05, \*\**p* < 0.01, \*\*\**p* < 0.001, \*\*\*\**p* < 0.0001. Abbreviations: UPDRS, Unified Parkinson’s Disease Rating Scale; ESS, Epworth Sleepiness Scale; RBDSQ, REM Sleep Behavior Disorder Screening Questionnaire; SCOPA-AUT, Scale for Outcomes in Parkinson’s Disease-Autonomic; HVLT-R, Hopkins Verbal Learning Test–Revised; BJLOT, Benton Judgment of Line Orientation test; LNS, Letter Number Sequencing test; MoCA, Montreal Cognitive Assessment.

### 2.2. Image acquisition

The Siemens 3T Trio or Verio scanners, manufactured by Siemens Medical Solutions in Malvern, PA, were used to obtain both structural and functional imaging data. The 3D T1-weighted MRI images were acquired using a magnetization-prepared rapid acquisition gradient echo sequence: Repetition time (TR) = 2300 ms, Echo time (TE) = 2.98 ms, Voxel size = 1 mm^3^, Slice thickness = 1.2 mm, twofold acceleration, and sagittal-oblique angulation. The DTI was acquired using the following specific parameters: TR = 8,400-8,800 ms, TE = 88 ms, Voxel size = 2 mm^3^, Slice thickness = 2 mm, 64 diffusion-sensitive gradient directions at b = 1,000 s/mm^2^ and one diffusion-unweighted (b0) image.

### 2.3. Imaging preprocessing

The preprocessing of DTI images in 146 PD patients was conducted using the FMRIB Software Library toolkit (FSL, https://fsl.fmrib.ox.ac.uk/fsl/fslwiki). In short, the DTI images were initially corrected for head motions, eddy-currents distortions, and artifacts. Afterwards, fractional anisotropy (FA), mean diffusivity (MD), axial diffusivity, and radial diffusivity were computed. Finally, the processed images were further reconstructed in the standard Montreal Neurological Institute (MNI) space to generate structural networks. The detailed preprocessing procedures of DTI images have been documented by our previous studies [6, 13].

### 2.4. Network construction

The structural networks were constructed using deterministic fiber tractography implemented in a free MATLAB toolkit, PANDA (http://www.nitrc.org/projects/panda/). Briefly, white matter fibers connecting all 90 cortical and subcortical nodes in the Automated Anatomical Labeling (AAL) atlas were generated using Fiber Assignment by Continuous Tracking (FACT) algorithm. The threshold for FA and fiber angle were 0.20 and 45°, respectively. The fiber numbers were used as edges for 90 x 90 structural network matrix of individual participant. Because structural network matrix constructed by FA and fiber numbers exhibited similar network topology [6, 25], we only analyzed structural network matrix based on fiber numbers in current study.

### 2.5. Graph-based network analysis

The GRETNA toolbox (https://www.nitrc.org/projects/gretna/) was used to compute the topological properties of structural network. Different network sparsity thresholds ranging from 0.05 to 0.50 with an interval of 0.05 were applied to calculate both global network metrics (global efficiency, local efficiency, and small-worldness metrics: clustering coefficient [Cp], characteristic path length [Lp], normalized clustering coefficient [γ], normalized characteristic path length [λ], and small worldness [σ]) and nodal network metrics (betweenness centrality, degree centrality, Cp, efficiency, local efficiency, and shortest path length). The area under curve (AUC) was calculated for both global and nodal network metrics. The detailed definitions for all network measurements calculated by our study have been documented by previous studies [26, 27].

### 2.6. Statistical analysis

#### 2.6.1. Evaluations of effects of SDMT scores on clinical variables

Two statistical methods were utilized to examine how SDMT scores affect clinical characteristics of PD patients. Firstly, the group differences of continuous clinical variables among 3 SDMT quartile groups were analyzed using One-way ANOVA test followed by Tukey’s post hoc test (Q1 group *vs* Q2-3 group *vs* Q4 group). The categorical variable (sex; Male = 1, Female = 2) was analyzed using χ^2^test. Secondly, to adjust the effects of confounding variables, the associations between SDMT scores and other clinical variables were analyzed with multivariate regression analysis with age, sex, years of education, and disease duration as covariates. *p* < 0.05 was considered statistically significant.

#### 2.6.2. Comparison of global network strength

The group differences of global network strength were analyzed using Network-Based Statistic (NBS, https://www.nitrc.org/projects/nbs/) toolbox. False discovery rate (FDR)-corrected *p* < 0.05 was considered statistically significant [28]. Age, sex, years of education, and disease duration were enrolled as covariates during NBS analysis.

#### 2.6.3. Comparison of network metrics

The group differences of global and nodal network metrics were analyzed using two-way ANOVA test followed by FDR corrections. FDR-corrected *p* < 0.05 was considered statistically significant.

#### 2.6.4. Association analysis between network metrics and clinical variables

The associations between clinical variables and network metrics were analyzed with Pearson correlation method and multivariate regression analysis with age, sex, years of education, and disease duration as covariates. FDR-corrected *p* < 0.05 was considered statistically significant.

#### 2.6.5. Mediation analysis

Mediation analysis was conducted using IBM SPSS Statistics Version 26. Age and network metrics were entered as independent variables during mediation analysis. The dependent variables were the scores of clinical assessments (UPDRS-III, total UPDRS, BJLOT, LNS and SDMT). The mediators were SDMT scores or graphical network metrics. The covariates including age, sex, disease duration, and years of education, were enrolled during the mediation analysis. *p* < 0.05 was considered statistically significant for mediation analysis.

## 3. Results

### 3.1. Group differences in clinical variables

As shown in Table S1, compared to Q4 group, Q1 group showed higher age (*p* < 0.0001), male dominance (*p* = 0.0364), and much lower years of education (*p* = 0.0035; Table S1). In addition, compared to Q4 group, Q2-3 group also showed higher age (*p* = 0.0008; Table S1). The disease duration was not statistically different among 3 quartile groups (Table S1). Compared to Q1 group, Q2-3 (*p* < 0.0001) and Q4 group (*p* < 0.0001) exhibited higher SDMT scores (Table S1). Compared to Q2-3 group and Q4 group, Q1 group displayed higher UPDRS-III scores (Fig. 1A), total UPDRS scores (Fig. 1B), ESS scores (Fig. 1C), RBDSQ scores (Fig. 1D), SCOPA-AUT scores (Fig. 1E), and lower scores of BJLOT (Fig. 1F), LNS (Fig. 1G), MoCA (Fig. 1H), Immediate Recall and Delayed Recall of HVLT-R (Fig. 1I-J).

Compared to Q2-3 and Q4 group, Q1 group also showed much lower SBRs in right caudate (Fig. S1A), left caudate (Fig. S1B), left putamen (Fig. S1C), right striatum (Fig. S1D), left striatum (Fig. S1E), bilateral caudate (Fig. S1F), bilateral putamen (Fig. S1G), and bilateral striatum (Fig. S1H).

### 3.2. Associations between SDMT scores and clinical variables

For demographic variables, age (β = –0.35, *p* < 0.0001), sex (β = 4.83, *p* = 0.0049; Male = 1, Female = 2), and years of education (β = 0.99, *p* = 0.0004) were significantly associated with SDMT scores while disease duration was not significantly associated with SDMT scores. For clinical assessments, as shown in Table 1, SDMT scores were negatively associated with scores of UPDRS-III (*p* = 0.0024), total UPDRS (*p* < 0.0001), ESS (*p* = 0.0006), RBDSQ (*p* = 0.0012), SCOPA-AUT (*p* = 0.0343), STAI (*p* = 0.0391), and positively associated with the scores of BJLOT (*p* = 0.0004), SFT (*p* < 0.0001), LNS (*p* < 0.0001), MoCA (*p* < 0.0001), Immediate Recall (*p* = 0.0005) and Delayed Recall (*p* = 0.0007) of HVLT-R.

**Table 1.** Associations between SDMT scores and clinical assessments.

| Clinical assessments |  |  |  |  |  |  |  |  |
| --- | --- | --- | --- | --- | --- | --- | --- | --- |
| Variables | HY stage | Tremor | Rigidity | UPDRS-III | UPDRS-Total | ESS | GDS | RBDSQ |
| Statistics |  |  |  |  |  |  |  |  |
| $\beta$ value | $\beta = -0.007$ | $\beta = -0.04$ | $\beta = -0.04$ | $\beta = -0.26$ | $\beta = -0.51$ | $\beta = -0.11$ | $\beta = -0.04$ | $\beta = -0.07$ |
| $p$-value | $p = 0.1408$ | $p = 0.2234$ | $p = 0.1574$ | $p = 0.0024$ | $p < 0.0001$ | $p = 0.0006$ | $p = 0.0877$ | $p = 0.0012$ |
| Variables | SCOPA-AUT | STAI | BJLOT | SFT | LNS | MoCA | HVLT-R<br>Immediate Recall | HVLT-R<br>Delayed Recall |
| Statistics |  |  |  |  |  |  |  |  |
| $\beta$ value | $\beta = -0.10$ | $\beta = -0.34$ | $\beta = 0.06$ | $\beta = 0.43$ | $\beta = 0.12$ | $\beta = 0.11$ | $\beta = 0.15$ | $\beta = 0.08$ |
| $p$-value | $p = 0.0343$ | $p = 0.0391$ | $p = 0.0004$ | $p < 0.0001$ | $p < 0.0001$ | $p < 0.0001$ | $p = 0.0005$ | $p = 0.0007$ |

The data were shown as the $\beta$ and $p$ -values derived from multivariate regression analysis with age, sex, years of education, and disease duration as covariates. The motor function examination was assessed in ON state. $p < 0.05$ was considered statistically significant. Abbreviations: HY, Hoehn & Yahr; UPDRS, Unified Parkinson's Disease Rating Scale; ESS, Epworth Sleepiness Scale; RBDSQ, REM Sleep Behavior Disorder Screening Questionnaire; GDS, Geriatric Depression Scale; SCOPA-AUT, Scale for Outcomes in Parkinson's Disease-Autonomic; SDMT, Symbol Digit Modalities Test; LNS, Letter Number Sequencing; SFT, Semantic Fluency Test Score; STAI, State-Trait Anxiety Inventory; BJLOT, Benton Judgement of Line Orientation; MoCA, Montreal Cognitive Assessment; HVLT-R, Hopkins Verbal Learning Test-Revised.

### 3.3. Group differences in brain network metrics

For global structural network metrics, compared to Q2-3 and Q4 group, Q1 group displayed lower global efficiency (FDR-corrected *p* < 0.05, sparsity range: 0.15∼0.50; Fig. 2A), local efficiency (FDR-corrected *p* < 0.05, sparsity range: 0.15∼0.50; Fig. 2B), and higher small-worldness Lp (FDR-corrected *p* < 0.05, sparsity range: 0.15∼0.50; Fig. 2C). Consistently, Q1 group showed lower AUC of global efficiency (FDR-corrected *p* < 0.05; Fig. 2D), AUC of local efficiency (FDR-corrected *p* < 0.05; Fig. 2E), and higher AUC of small-worldness Lp (FDR-corrected *p* < 0.05; Fig. 2F) compared to Q2-3 and Q4 group. For nodal betweenness centrality in structural network, Q1 group showed both lower nodal betweenness centrality (FDR-corrected *p* < 0.05 in right superior frontal gyrus, right superior occipital gyrus, right precuneus; Fig. 3A) and higher nodal betweenness centrality (FDR-corrected *p* < 0.05 in bilateral putamen, right superior temporal gyrus, right middle temporal gyrus; Fig. 3A) compared to Q4 group. Importantly, compared to Q2-3 and Q4 group, Q1 group exhibited lower nodal degree centrality (FDR-corrected *p* < 0.05; Fig. 3B), nodal Cp (FDR-corrected *p* < 0.05; Fig. 3C), nodal efficiency (FDR-corrected *p* < 0.05; Fig. 3D), nodal local efficiency (FDR-corrected *p* < 0.05; Fig. 3E), and higher nodal shortest path length (FDR-corrected *p* < 0.05; Fig. 3F).

**Fig. 2.**
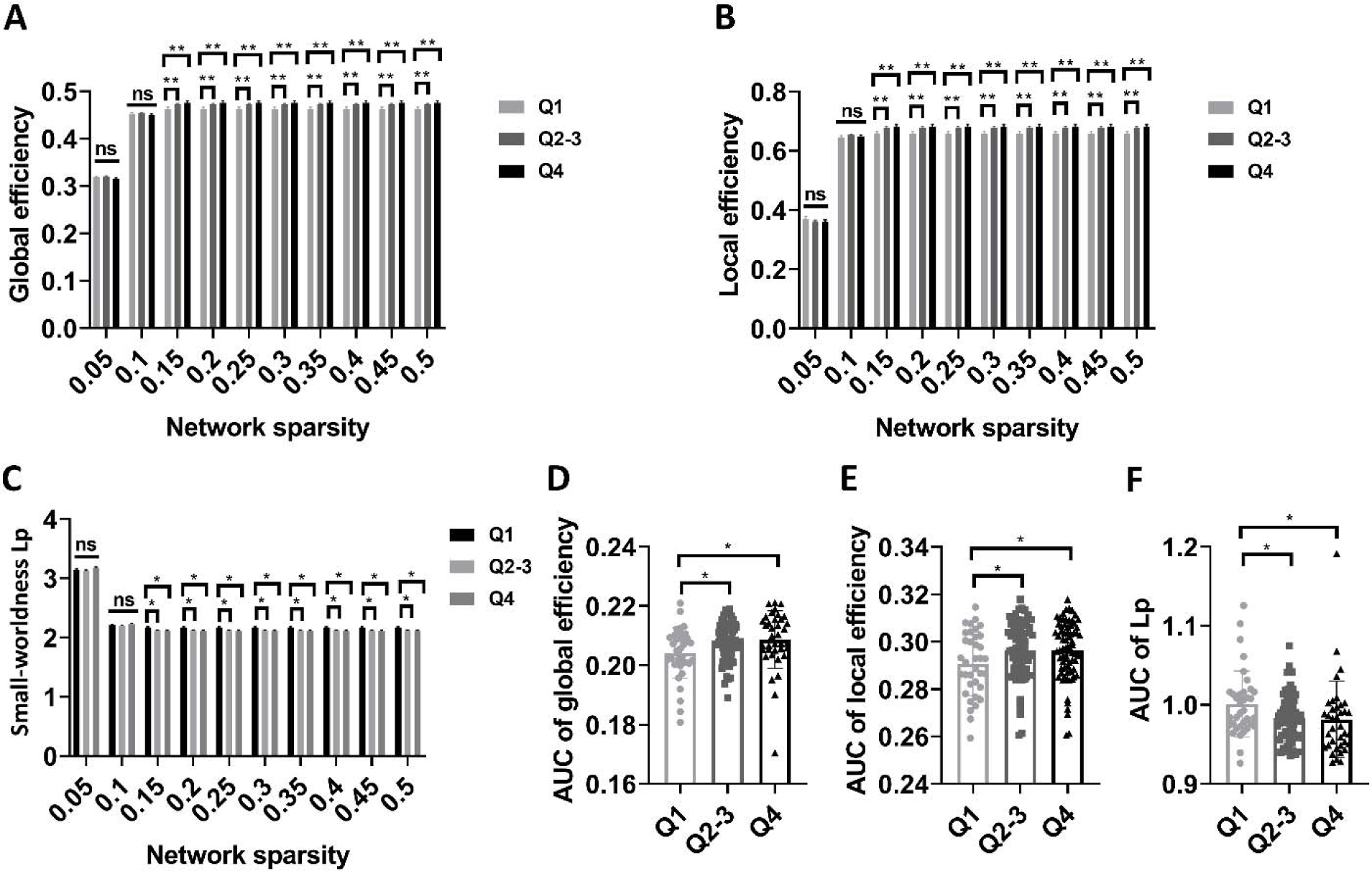
Group differences in global network metrics of structural network. (A-C) Group differences of global efficiency, local efficiency, and small-worldness Lp at different sparsity threshold (FDR-corrected *p* < 0.05; Two-way ANOVA test). (D-F) Group differences in AUCs of global efficiency, local efficiency, and small-worldness Lp (FDR-corrected *p* < 0.05; One-way ANOVA test). Two-way ANOVA test with FDR correction was performed to compare global efficiency, local efficiency, and small-worldness Lp at different sparsity threshold. One-way ANOVA test followed by FDR corrections was conducted to compare AUCs of global efficiency, local efficiency, and small-worldness Lp. FDR-corrected *p* < 0.05 was considered statistically significant. \**p* < 0.05, \*\**p* < 0.01. Abbreviations: AUC, Area under curve; Lp, Characteristic path length.

**Fig. 3.**
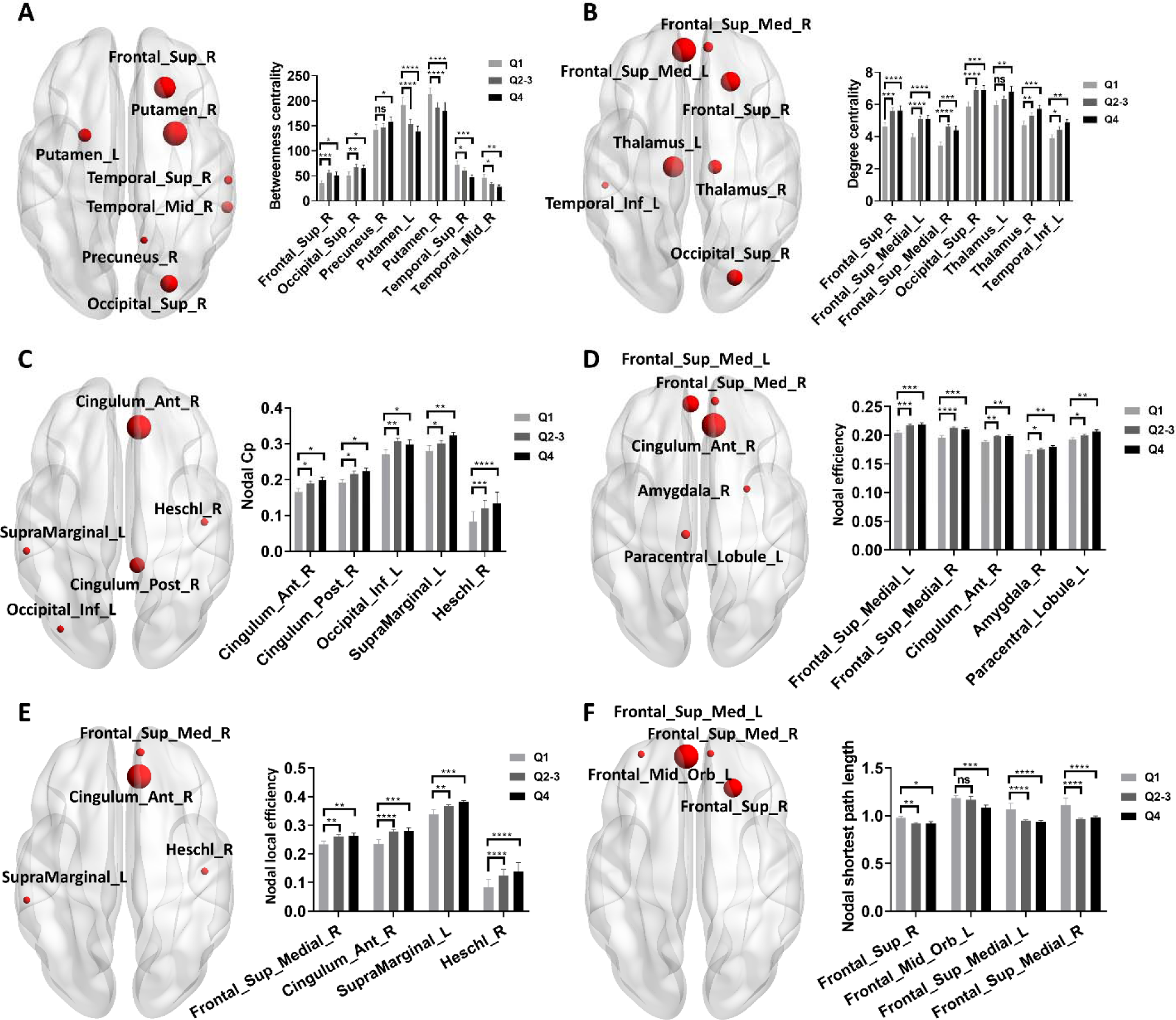
Group differences in nodal network metrics of structural network. (A-F) Group differences of nodal betweenness centrality, nodal degree centrality, nodal Cp, nodal efficiency, nodal local efficiency, and nodal shortest path length (FDR-corrected *p* < 0.05; Two-way ANOVA test). Two-way ANOVA test with FDR corrections was performed. FDR-corrected *p* < 0.05 was considered statistically significant. \**p* < 0.05, \*\**p* < 0.01, \*\*\**p* < 0.001, \*\*\*\**p* < 0.0001. Abbreviations: Cp, Clustering coefficient.

### 3.4. Associations between graphical network metrics and clinical assessments

Among above differential network metrics shown in Figure 2 and 3, we identified some network metrics were significantly associated with clinical assessments. For example, nodal betweenness centrality of right middle temporal gyrus was negatively associated with scores of LNS, SDMT, SFT, MoCA, and Immediate Recall of HVLT-R (FDR-corrected *p* < 0.05 in both Pearson correlation and multivariate regression analysis; Fig. S2A). Nodal degree centrality of right superior occipital gyrus was positively associated with scores of BJLOT and MoCA scores (FDR-corrected *p* < 0.05 in both Pearson correlation and multivariate regression analysis; Fig. S2B). Nodal efficiency of right anterior cingulate cortex was negatively associated with UPDRS-III scores and total UDPRS scores (FDR-corrected *p* < 0.05 in both Pearson correlation and multivariate regression analysis; Fig. S2C). Additionally, nodal efficiency of right anterior cingulate cortex was positively associated with scores of SFT, SDMT, BJLOT, and MoCA (FDR-corrected *p* < 0.05 in both Pearson correlation and multivariate regression analysis; Fig. S2D).

### 3.5. Mediation analysis

In a recent study, we demonstrated that age was an essential demographic factor significantly shaping clinical features of PD patients and global structural network metrics mediated the effects of age on SFT and SDMT scores [13]. In this study, we examined whether nodal (local) network metrics also mediated the effects of age on clinical assessments. As shown in Figure 4, we found nodal betweenness centrality of right middle temporal gyrus mediated the effects of age on LNS scores (Fig. 4A). Degree centrality of right superior occipital gyrus and nodal efficiency of right anterior cingulate cortex mediated the effects of age on BJLOT scores (Fig. 4B-C). Nodal betweenness centrality of right middle temporal gyrus, nodal efficiency of right anterior cingulate cortex, and nodal efficiency of left paracentral lobule mediated the effects of age on SDMT scores (Fig. 4D-F).

**Fig. 4.**
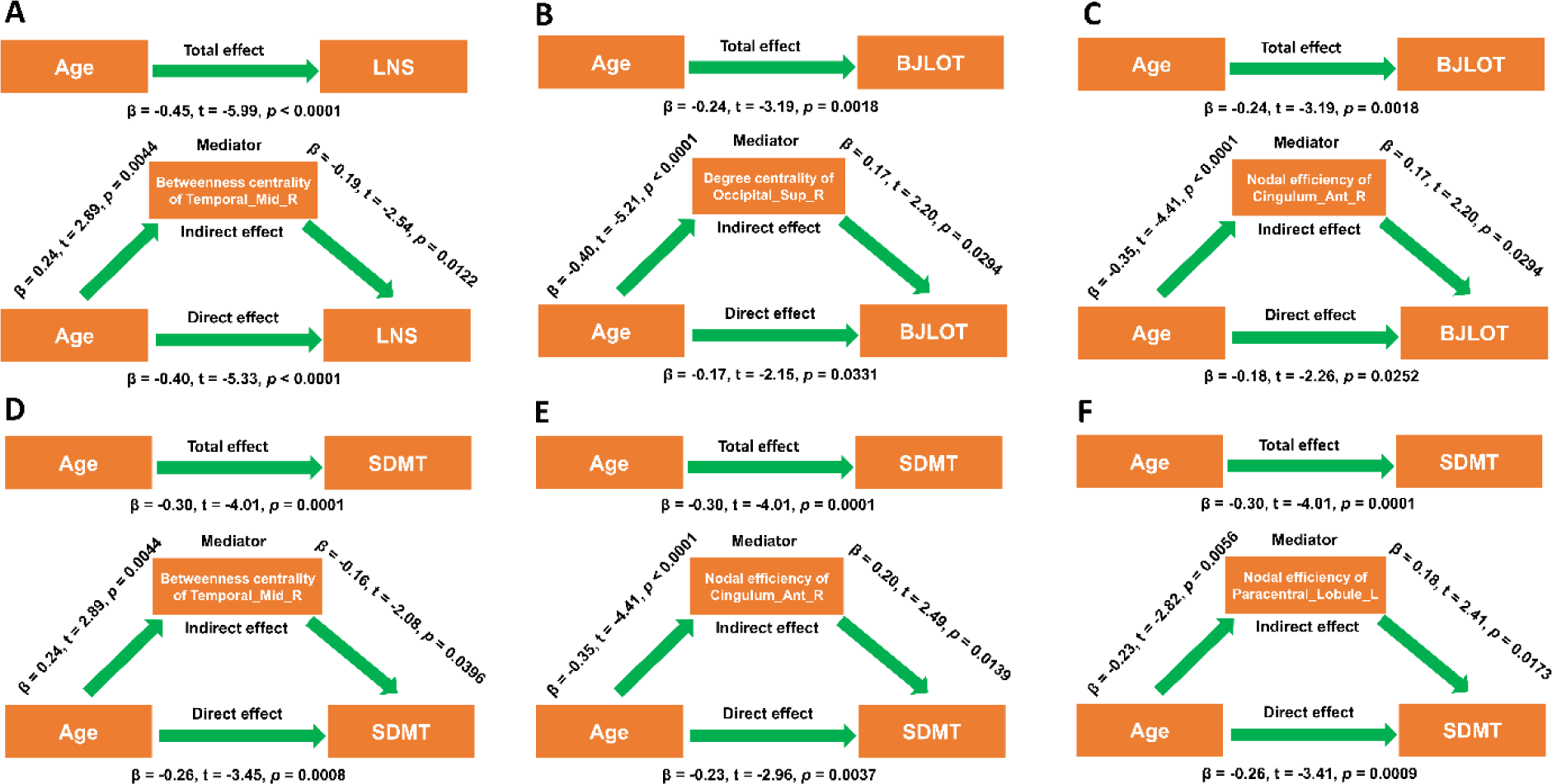
Structural network metrics mediated the effects of age on cognitive decline. (A) Nodal betweenness centrality of right middle temporal gyrus mediated the effects of age on LNS scores. (B-C) Degree centrality of right superior occipital gyrus and nodal efficiency of right anterior cingulate cortex mediated the effects of age on BJLOT scores. (D-F) Nodal betweenness centrality of right middle temporal gyrus, nodal efficiency of right anterior cingulate cortex, and nodal efficiency of left paracentral lobule mediated the effects of age on SDMT scores. During the mediation analysis, age, sex, disease duration, and years of education were included as covariates. *p* < 0.05 was considered statistically significant. Abbreviations: BJLOT, Benton Judgment of Line Orientation test; LNS, Letter Number Sequencing test; SDMT, Symbol Digit Modalities Test.

In Supplementary Figure 2, we found nodal efficiency of right anterior cingulate cortex was negatively associated with UPDRS-III scores and total UDPRS scores, here, we further demonstrated that nodal efficiency of right anterior cingulate cortex may shape UPDRS-III scores and total UDPRS scores by modulating cognitive function, which was measured by SDMT (Fig. S3A-B).

## 4. Discussion

In this study, we found PD patients with better cognition exhibited milder clinical symptoms compared to PD patients with worse cognitive function. Consistently, they also displayed higher striatum SBRs, indicating relatively milder dopaminergic neurodegeneration. According to previous literature, this is the first study showing that SDMT-measured cognitive function is an important predictor of disease severity in PD patients. With graphical analysis, we revealed that PD patients with better cognition showed relatively superior structural network topology at both global and nodal levels compared to PD patients with worse cognitive function, indicating SDMT was a powerful instrument specifically measuring the global and local information transfer efficiency of white matter network. Using mediation analysis, we further demonstrated that local structural topological metrics mediated the effects of age on cognitive decline of PD patients. Taken together, our findings suggest that SDMT is an excellent tool to monitor disease severity and specifically reflects the information transfer efficiency of structural network. In addition, our results also demonstrate that local topology of structural network is causally associated with age-dependent cognitive decline in PD patients, which deepens our understanding about the neural mechanisms underlying cognitive impairment of PD patients.

### 4.1. Cognition and clinical symptoms of PD patients

The SDMT is a widely used neuropsychological instrument assessing multiple cognitive domains, such as divided attention, information processing speed, visuospatial ability, and memory [21]. It has been used for the evaluation of information processing speed in multiple sclerosis [29, 30]. Basically, lexical access speed, memory, and information processing speed all independently contribute to the performance of SDMT [30]. In this study, we found SDMT was significantly and positively associated with the performance of other cognitive scales, such as BJLOT, LNS, SFT, and MoCA, which also suggested SDMT could measure multiple cognitive processes in PD. According to previous studies, cognitive impairment in PD was associated with a variety of risk factors, such as age, education, motor severity, depression, anxiety, and excessive daytime sleepiness (EDS) [10–13, 31, 32]. Here, we showed older age and lower education level were associated with worse cognitive function in PD patients, which was consistent with our recent findings [13]. Therefore, age and education were key demographics that significantly shaped cognitive function of PD patients. We found male patients were more likely to exhibit cognitive decline than female patients, which was supported by our recent study showing that female sex was associated with better cognition in PD patients [13]. We found upper quartile group and interquartile group exhibited lower UPDRS-III scores compared to lower quartile group, indicating that better cognition corresponded to milder motor severity. This was demonstrated by our association analysis showing SDMT scores were negatively associated with UPDRS-III scores. These findings indicated that cognitive function may modify the motor severity of PD patients, which was supported by a recent study showing that freezing of gait was associated with cognitive impairment in PD patients [33]. EDS was one of the most common non-motor symptoms in PD patients [34]. It was associated with both worse motor and non-motor manifestations [34]. Especially, EDS was associated with the impairment of executive control and processing speed in PD patients [31, 32]. Consistent with these findings, here, we also found cognitive function was negatively associated with ESS scores and better cognitive level predicted lower ESS scores. We found SDMT scores were negatively associated with RBDSQ scores, which was consistent with the results reported by previous studies showing that PD patients with RBD exhibited impaired global cognition, verbal memory, and attention compared to PD patients without RBD [35, 36]. We found SDMT scores were negatively associated with SCOPA-AUT scores, which was in agreement with recent studies showing that autonomic dysfunction was associated with cognitive impairment in PD patients [37, 38].

Anxiety was also associated with the impairment of multiple cognitive domains, spanning attention, executive function, working memory, and language in PD patients [39]. Additionally, anxiety was found to be associated with the development of mild cognitive impairment in PD patients [40]. These results were consistent with our finding that SDMT scores were negatively associated with STAI scores. Importantly, we found better cognitive function was also associated with relatively milder dopaminergic neurodegeneration measured by stratum SBRs, which suggested that cognitive function evaluated by SDMT could be a potential predictor of striatal dopamine transporter availability. Taken together, cognitive function measured by SDMT was a potential predictor for disease severity and progression in PD, which deserved to be further investigated in future studies.

### 4.2. Structural network metrics and cognition in PD patients

As mentioned above, we found cognitive function in PD patients was significantly associated with both motor and non-motor manifestations. However, the neural mechanisms underlying cognitive impairment or cognitive maintenance in PD patients remain unclear. fMRI has been used to investigate the neural correlates of cognitive impairment in PD patients and both structural and functional metrics were found to be associated with cognitive decline in PD patients [13, 14]. SDMT is preferential for the assessment of white matter-related cognitive function, such as information processing speed and verbal memory [41]. During SDMT, white matter activation can be detected in multiple white matter tracts, especially corpus callosum (anterior and/or posterior) or internal capsule (left and/or right) [41]. Recently, we found global network metrics in structural network, such as global efficiency and local efficiency, were positively associated with SDMT scores [13], which indicated that SDMT performance reflected the information transfer efficiency of global white matter structure in PD patients. Consistently, in current study, we found PD patients with better cognition exhibited higher global efficiency and local efficiency in multiple sparsity thresholds compared to PD patients with worse cognition. Therefore, we concluded that SDMT was an excellent instrument to measure both structural network integrity and information processing efficiency along white matter tracts [13, 30, 41]. In a recent study, we found the global network properties of white matter network were significantly associated with cognitive function [13], nevertheless, whether local/nodal network metrics correlated with cognitive function remain unclear. In this study, we revealed PD patients with better cognition displayed higher nodal degree centrality, nodal Cp, nodal efficiency, nodal local efficiency, and lower nodal shortest path length compared to PD patients with worse cognitive function. Importantly, all of these nodal metrics exhibited does-dependent changes according to the SDMT quartiles, which suggested that cognitive function in PD patients was determined by a wide range of brain nodes and nodal network metrics, such as nodal efficiency and nodal local efficiency, were also powerful predictors of cognitive function in PD patients. Different from other nodal network metrics, nodal betweenness centrality showed bidirectional changes across different SDMT quartiles, indicating that nodal betweenness centrality did not always change in the same direction as the cognitive function.

We revealed multiple nodal network metrics were significantly associated with cognitive function of PD patients. For instance, betweenness centrality of right middle temporal gyrus was negatively associated with multiple cognitive domains in PD patients, which suggested that betweenness centrality of right middle temporal gyrus was a reliable predictor of cognitive function in PD patients. Therefore, we hypothesized that the changes of structure and function of right middle temporal gyrus might be associated with cognitive function in PD patients. In agreement with this hypothesis, PD patients with cognitive impairment exhibited lower gray matter volume in temporal lobe compared to patients without cognitive impairment [42]. Moreover, Chiang *et al*. (2018) revealed that disruption of bilateral medial temporal lobes contributed to cognitive decline in PD patients [43]. Importantly, using mediation analysis, we further demonstrated that betweenness centrality of right middle temporal gyrus mediated the effects of age on LNS scores and SDMT scores, suggested that right middle temporal gyrus was causally associated with cognitive impairment in PD patients. We found degree centrality of right superior occipital gyrus were significantly associated with scores of BJLOT and MoCA, indicating that degree centrality of right superior occipital gyrus was also potential imaging predictor of cognitive function in PD patients. According to previous studies, right superior occipital gyrus has been demonstrated to be a key region involved in cognitive impairment in a variety of diseases. For instance, the cortical volume of the right superior occipital gyrus was found to be associated with the conversion of amnestic mild cognitive impairment to Alzheimer’s disease in patients with type 2 diabetes [44]. In addition, the regional homogeneity (ReHo) of right superior occipital gyrus was positively correlated with speed of processing and social cognition in schizophrenia [45]. In PD patients, functional connectivity between right superior occipital gyrus and left superior parietal gyrus was significantly associated with MoCA and visuospatial skills/executive function scores while functional connectivity between right superior occipital gyrus and left calcarine was significantly associated with delayed memory scores in PD patients with RBD [46]. Using mediation analysis, we further demonstrated that degree centrality of right superior occipital gyrus mediated the effects of age on BJLOT scores, suggesting that right superior occipital gyrus was an essential brain region significantly correlated with cognitive function in PD patients, especially visuospatial capacity. We found nodal efficiency of right anterior cingulate cortex was associated with the performance of multiple cognitive scales, such as SFT, SDMT, BJLOT, and MoCA, indicating that right anterior cingulate cortex may be also associated with cognitive impairment in PD patients. Indeed, we demonstrated that nodal efficiency of right anterior cingulate cortex mediated the effects of age on BJLOT scores and SDMT scores. The role of right anterior cingulate cortex in cognitive function have been shown by a series of studies. For example, decreased functional connectivity between the left posterior cingulate cortex and right anterior cingulate cortex was found to be associated with longer completion time in Wisconsin Card Sorting Test in bipolar disorder patients [47]. It was also shown that multiple sclerosis patients with worsened cognition had more severe gray matter atrophy in right anterior cingulate cortex compared to cognitively stable multiple sclerosis patients [48]. Additionally, ReHo in right anterior cingulate cortex has been found to be negatively correlated with working memory in schizophrenia patients [45]. In PD patients, blood perfusion was significantly decreased in right anterior cingulate cortex compared with the normal controls [49]. Furthermore, amplitude of low-frequency fluctuation in right anterior cingulate cortex was positively correlated with freezing of gait [50], which was causally associated with cognitive impairment in PD patients [4, 33]. Taken together, these findings suggested that right anterior cingulate cortex might play an essential role in the development of cognitive impairment in PD patients.

### 4.3. Structural network metrics and motor impairment of PD patients

In this study, we found nodal efficiency of right anterior cingulate cortex was also associated with UPDRS-III scores and total UPDRS scores, which indicated that right anterior cingulate cortex may be also involved in motor impairment in PD patients. With mediation analysis, we further demonstrated that cognitive function may mediated the effects of right anterior cingulate cortex on UPDRS-III scores and total UPDRS scores. Therefore, we hypothesized that cognitive impairment contributed to motor decline in PD patients [51]. In agreement with this hypothesis, a previous study revealed cognitive impairment was associated with higher scores of UPDRS, bradykinesia, and rigidity [52]. In addition, freezing of gait in PD patients was also significantly associated with cognitive impairment [4, 33]. Specifically, it has been shown that executive function and attention was significantly associated with bradykinesia and rigidity, while visuospatial function was associated with bradykinesia and tremor [53]. Moreover, PD patients with mild cognitive impairment presented a more severe bradykinesia score than PD patients without mild cognitive impairment [54]. Thus, these findings suggested that cognitive decline exacerbated motor impairment in PD patients. In this study, we found nodal efficiency of right anterior cingulate cortex was negatively associated with UPDRS-III and total UPDRS scores, which indicated that it was a potential predictor of motor progression in PD patients. Indeed, we found nodal efficiency of right anterior cingulate cortex was also negatively associated with HY stages (r = –0.21, *p* = 0.0137) in PD patients. Although our study supported right anterior cingulate cortex was involved in motor impairment in PD patients as previously reported [50], its specific roles in motor decline of PD patients remained largely unclear and were required to be further investigated.

### 4.4. Strengths and limitations of this study

In this study, we found PD patients with better cognition measured by SDMT displayed milder motor and non-motor symptoms compared to patients with worse cognition, which suggested cognitive function measured by SDMT was a potential predictor of disease progression in PD patients. We found PD patients with better cognition exhibited higher information transfer efficiency both at global and local levels in structural network, which indicated that structural network topology was a key determinator of cognitive function in PD patients. This was further supported by our results that structural topological metrics were significantly associated with cognitive function in PD patients [13]. With mediation analysis, we demonstrated that structural topological metrics mediated the effects of age on cognitive function of PD patients, which provided new evidence to our recent proposal that age significantly shaped cognitive function in PD patients by regulating structural network topology [13]. Specifically, we identified some brain regions, such as right middle temporal gyrus, right superior occipital gyrus, and right anterior cingulate gyrus, may played an important role in the development of cognitive decline in PD patients. Importantly, we revealed that cognitive function modified motor symptoms in PD patients, which emphasized the role of cognitive impairment in motor decline of PD patients. Because our findings mainly come from cross-section analysis, the major findings were required to be validated in future longitudinal studies.

To conclude, SDMT is an excellent instrument to monitor disease severity and to represent information transfer efficiency of structural network in PD patients. Additionally, local structural topological metrics were significantly associated with both cognitive and motor impairment of PD patients.

## Author contributions

Zhichun Chen, Conceptualization, Formal analysis, Visualization, Methodology, Writing, review and editing; Guanglu Li, Data curation, Formal analysis, Visualization; Liche Zhou, Data curation, Formal analysis, Investigation; Lina Zhang, Formal analysis, Investigation, Methodology; Li Yang, Supervision, Writing, review and editing; Jun Liu, Conceptualization, Supervision, Funding acquisition, Writing, review, and editing.

## Acknowledgments

Data used in the preparation of this article were obtained from the Parkinson’s Progression Markers Initiative (PPMI) database (www.ppmiinfo.org/data). We thank the share of PPMI data by all the PPMI study investigators. PPMI – a public-private partnership – is funded by the Michael J. Fox Foundation for Parkinson’s Research and funding partners, which can be found at www.ppmiinfo.org/fundingpartners.

## Data availability

All the raw data used in the preparation of this Article were downloaded from PPMI database (www.ppmi-info.org/data).All data produced in the present study are available upon reasonable request to the authors.

## Supporting Information

Additional supporting information may be found online in the Supporting Information section at the end of the article.

