## Supplementary Material for "Structural network topology correlates with cognitive impairment in Parkinson’s disease"

**Supplementary Materials**

| Clinical variable | Q1 group  (n = 36) | Q2-3 group  (n = 73) | Q4 group  (n = 37) | Q1 *vs* Q2-3 | Q1 *vs* Q4 | Q2-3 *vs* Q4 |
| --- | --- | --- | --- | --- | --- | --- |
| Age, years | 65.66 ± 7.99 | 61.95 ± 8.82 | 55.55 ± 8.38 | *p* = 0.0848 | ***p* < 0.0001** | ***p* = 0.0008** |
| Sex (Male/Female) | 27/9 | 47/26 | 19/18 | *p* = 0.2642 | ***p* = 0.0364** | *p* = 0.1874 |
| Education, years | 14.14 ± 3.10 | 15.26 ± 2.92 | 16.38 ± 2.65 | *p* = 0.1429 | ***p* = 0.0035** | *p* = 0.1395 |
| Disease duration, years | 2.43 ± 3.40 | 1.86 ± 1.30 | 2.02 ± 1.95 | *p* = 0.3999 | *p* = 0.6940 | *p* = 0.9305 |
| SDMT scores | 26.25 ± 6.42 | 40.85 ± 3.81 | 53.46 ± 5.77 | ***p* < 0.0001** | ***p* < 0.0001** | ***p* < 0.0001** |

**Table S1** The demographic and clinical data for each quartile group.

The data were shown as the mean ± standard deviation (SD). One-way ANOVA followed by Tukey’s post hoc test (Q1 group *vs* Q2-3 group *vs* Q4 group) were utilized to compare continuous variables. χ^2^ test was used to compare categorical variable (Sex). *p* < 0.05 was considered statistically significant. The bold values indicate statistical significance. Abbreviations: SDMT, Symbol Digit Modalities Test.

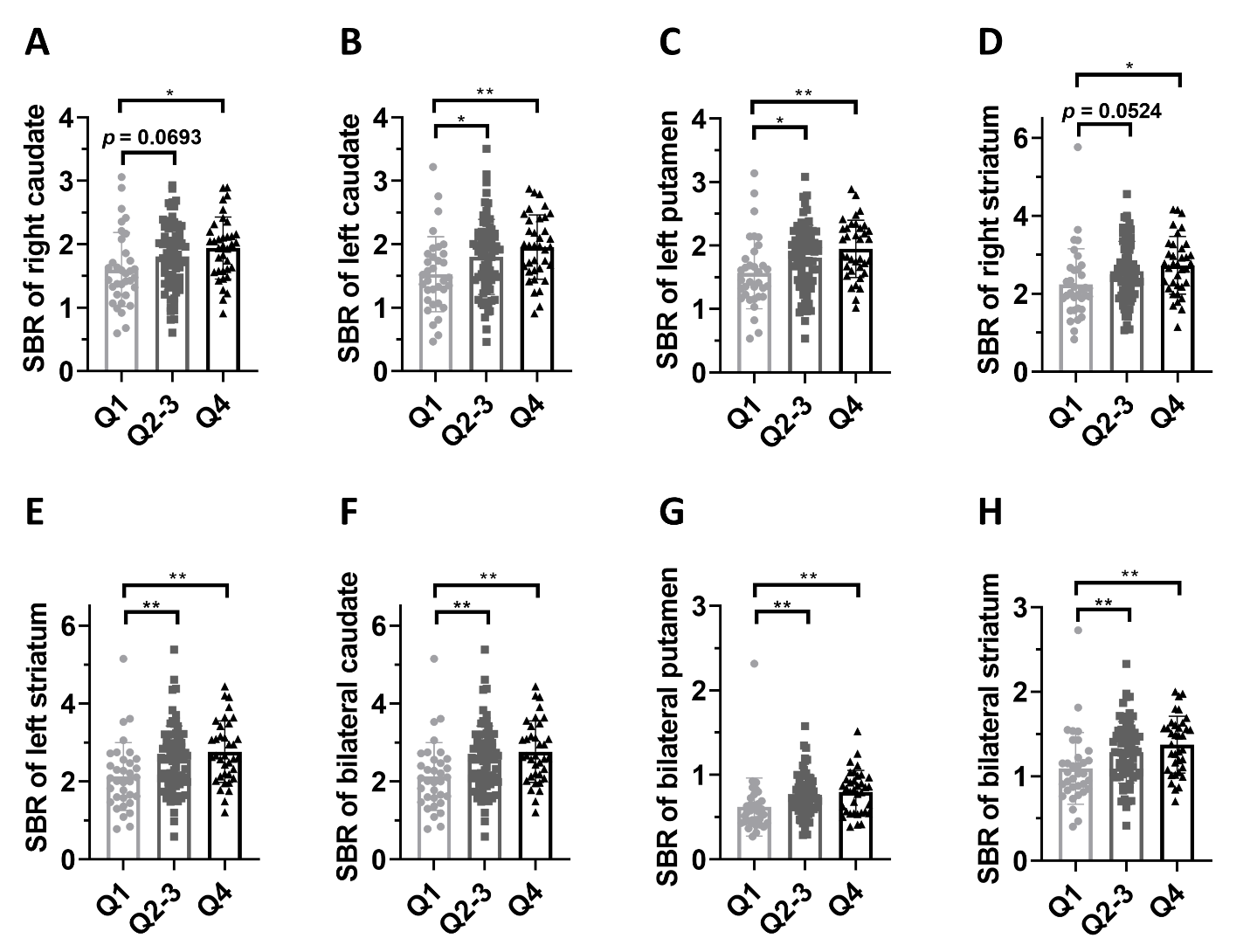

**Supplementary Fig. 1. Group differences in striatum SBRs.** (A-F) Group differences in SBRs of right caudate (A), left caudate (B), left putamen (C), right stratum (D), left stratum (E), bilateral caudate (F), bilateral putamen (G), and bilateral striatum (H). One-way ANOVA followed by Tukey’s post hoc test (Q1 group *vs* Q2-3 group *vs* Q4 group) were conducted to compare clinical variables. *p* < 0.05 was considered statistically significant. **p* < 0.05, ***p* < 0.01. Abbreviations: SBR, striatal binding ratio.

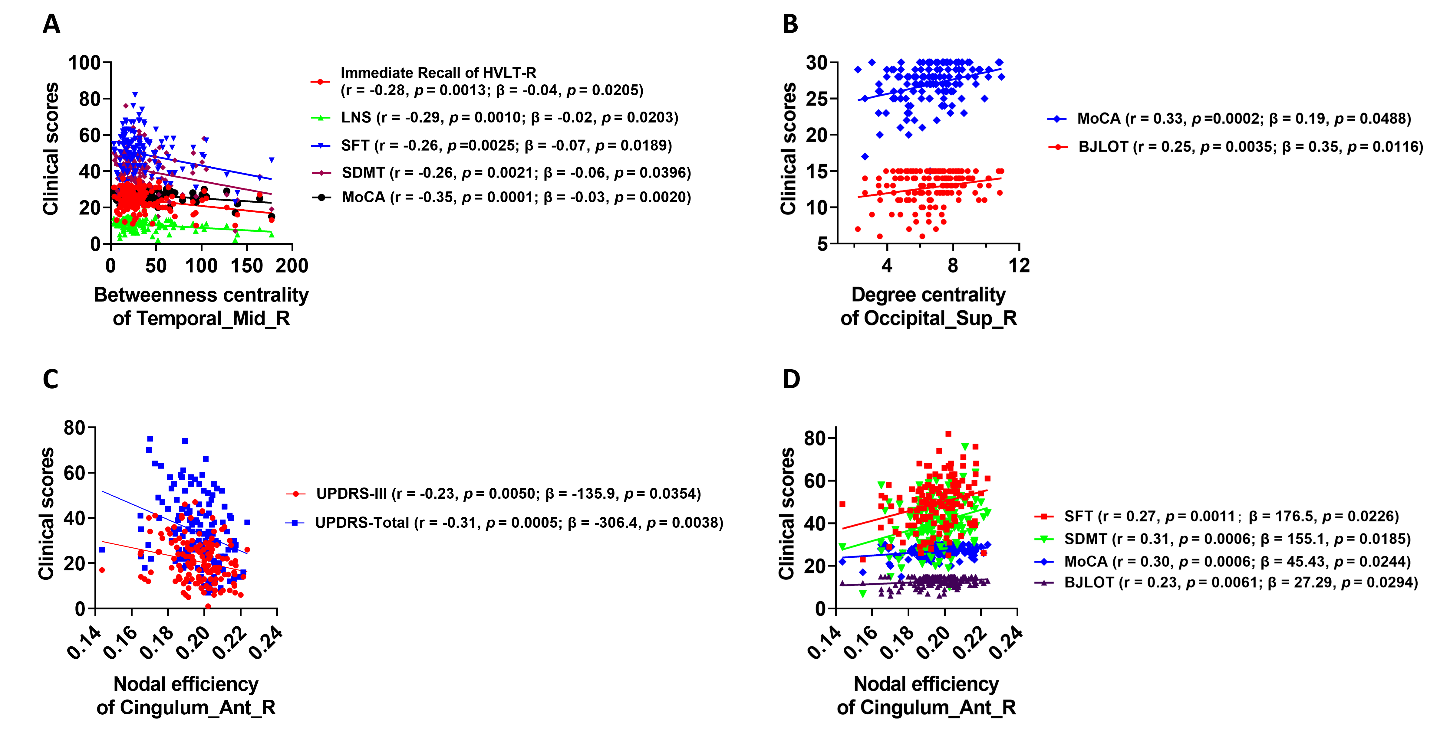

**Supplementary Fig. 2.** **Associations between nodal network metrics and clinical assessments.** (A) Nodal betweenness centrality of right middle temporal gyrus was negatively scores of LNS, SDMT, SFT, MoCA, and Immediate Recall of HVLT-R (FDR-corrected *p* < 0.05 in both Pearson correlation and multivariate regression analysis). (B) Nodal degree centrality of right superior occipital gyrus was positively associated with scores of MoCA and BJLOT scores (FDR-corrected *p* < 0.05 in both Pearson correlation and multivariate regression analysis). (C) Nodal efficiency of right anterior cingulate cortex was negatively associated with UPDRS-III scores and total UDPRS scores (FDR-corrected *p* < 0.05 in both Pearson correlation and multivariate regression analysis). (D) Nodal efficiency of right anterior cingulate cortex was positively associated with scores of SFT, SDMT, BJLOT, and MoCA (FDR-corrected *p* < 0.05 in both Pearson correlation and multivariate regression analysis). The association analysis between graphical network metrics and clinical assessments was conducted by both Pearson correlation method and multivariate regression analysis with age, sex, years of education, and disease duration as covariates. FDR-corrected *p* < 0.05 was considered statistically significant. Abbreviations: HVLT-R, Hopkins Verbal Learning Test–Revised; BJLOT, Benton Judgment of Line Orientation test; SFT, Semantic Fluency Test; SDMT, Symbol Digit Modalities Test; LNS, Letter Number Sequencing test; MoCA, Montreal Cognitive Assessment; UPDRS, Unified Parkinson’s Disease Rating Scale.

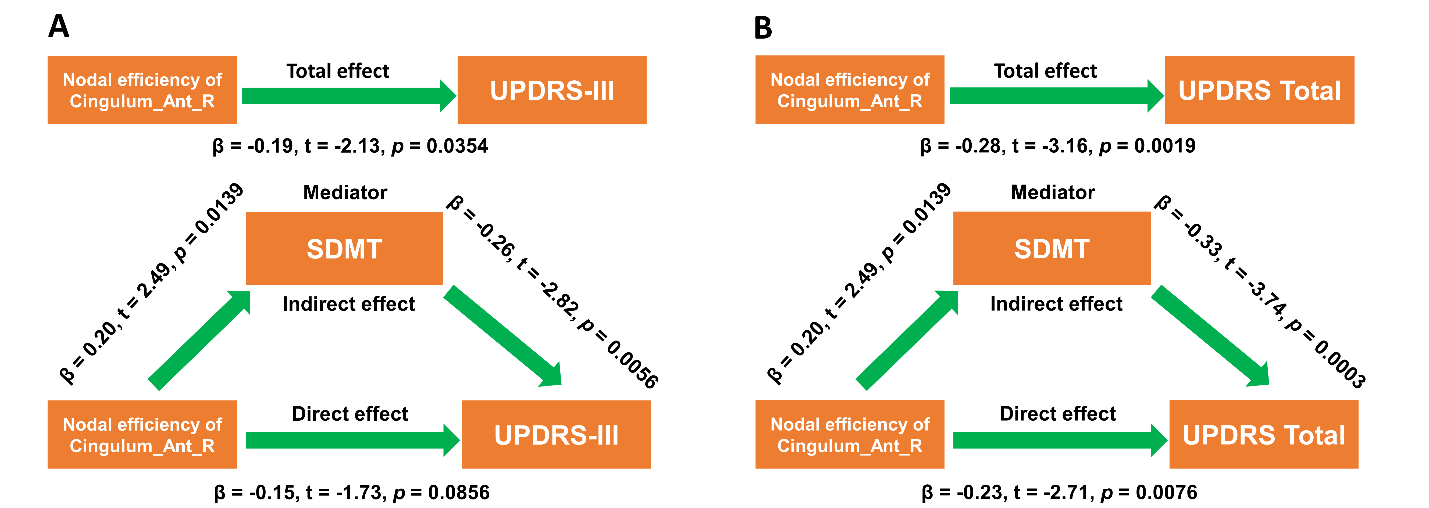

**Supplementary Fig. 3. Cognitive function mediated the negative associations between nodal network metrics and UPDRS-III scores or total UPDRS scores.** (A-B) Cognitive function mediated the effects of nodal efficiency in right anterior cingulate cortex on UPDRS-III scores (A) and total UDPRS scores (B). During the mediation analysis, age, sex, disease duration, and years of education were included as covariates. *p* < 0.05 was considered statistically significant. Abbreviations: SDMT, Symbol Digit Modalities Test; UPDRS, Unified Parkinson’s Disease Rating Scale.
